# Building a Vertically-Integrated Genomic Learning Health System: The Colorado Center for Personalized Medicine Biobank

**DOI:** 10.1101/2022.06.09.22276222

**Authors:** Laura K Wiley, Jonathan A Shortt, Emily R Roberts, Jan Lowery, Elizabeth Kudron, Meng Lin, David A Mayer, Melissa P Wilson, Tonya M Brunetti, Sameer Chavan, Tzu L Phang, Nikita Pozdeyev, Joseph Lesny, Stephen J Wicks, Ethan Moore, Joshua L Morgenstern, Alanna N Roff, Elise L Shalowitz, Adrian Stewart, Cole Williams, Michelle N Edelmann, Madelyne Hull, J. Tacker Patton, Lisen Axell, Lisa Ku, Yee Ming Lee, Jean Jirikowic, Anna Tanaka, Emily Todd, Sarah White, Brett Peterson, Emily Hearst, Richard Zane, Casey S Greene, Rasika Mathias, Marilyn Coors, Matthew RG Taylor, Debashis Ghosh, Michael G Kahn, Ian M Brooks, Christina L Aquilante, David Kao, Nicholas Rafaels, Kristy Crooks, Steve Hess, Kathleen C Barnes, Christopher R Gignoux, the Colorado Center for Personalized Medicine

## Abstract

Precision medicine initiatives across the globe have led to a revolution of repositories linking large-scale genomic data with electronic health records, enabling genomic analyses across the entire phenome. Many of these initiatives focus solely on research insights, leading to limited direct benefit to patients. We describe the Biobank at the Colorado Center for Personalized Medicine (CCPM Biobank) that was jointly developed by the University of Colorado Anschutz Medical Campus and UCHealth to serve as a unique, dual-purpose research and clinical resource accelerating personalized medicine. This living resource currently has over 200,000 patients with ongoing recruitment. We highlight the clinical, laboratory, regulatory, and HIPAA-compliant informatics infrastructure along with our stakeholder engagement, consent, recontact, and participant engagement strategies. We characterize aspects of genetic and geographic diversity unique to the Rocky Mountain Region, the primary catchment area for CCPM Biobank participants. We leverage linked health and demographic information of the CCPM Biobank participant population to demonstrate the utility of the CCPM Biobank to replicate complex trait associations in the first 33,674 genotyped patients across multiple disease domains. Finally, we describe our current efforts towards return of clinical genetic test results including high-impact pathogenic variants and pharmacogenetic information, and our broader goals as the CCPM Biobank continues to grow. Bringing clinical and research interests together fosters unique clinical and translational questions that can be addressed from the large EHR-linked CCPM Biobank resource within a HIPAA and CLIA-certified environment.

## INTRODUCTION

Population-scale biobanks increasingly provide opportunities to understand patterns of health and disease within and across populations, discover the factors that drive these patterns, and enable advances in healthcare.^1–4^ The Biobank at the Colorado Center for Personalized Medicine (CCPM Biobank) was jointly developed by the University of Colorado Anschutz Medical Campus and UCHealth to serve as a unique, dual-purpose research and clinical resource accelerating personalized medicine. The CCPM Biobank has broad objectives to collect, store, and generate data on biological samples within a clinically certified environment. The goals of these activities are to support scientific discovery, collaboration with partners, and return of actionable clinical results. As a resource comprising electronic health records (EHR), genotype data, and other integrated data sources (e.g., geocoded data and survey data), the CCPM Biobank is available to the University of Colorado (CU) community for discovery research and pragmatic clinical trials across a range of both acute and chronic conditions. From its inception, the CCPM Biobank was designed as a translational medicine initiative that delivers biobank-fueled innovations back to participants and their healthcare providers to inform healthcare decisions.

In the context of the ongoing revolution of personalized medicine initiatives worldwide, the CCPM Biobank fills a unique niche with diverse data from the Rocky Mountain Region. UCHealth serves both a highly urban population in the Denver, Colorado Springs, and Fort Collins metropolitan areas as well as numerous rural populations in the intermountain west and western great plains regions of the United States. UCHealth is comprised of 679 clinics and 13 hospitals, including a quaternary care facility and academic medical headquarters at the Anschutz Medical Campus in Aurora, Colorado. UCHealth has served over 6 million patients in the last 10 years including approximately 2 million active patients seen within the past 2 years. UCHealth patients enrolled in the CCPM Biobank receive care for a broad range of conditions and come from diverse and often unique populations with respect to medical conditions, socioeconomic status, geographic context, and cultural norms. In particular, our catchment includes many individuals living in rural and/or high altitude settings that are rare in other biobanks. Their participation enables unprecedented insights into interactions between genetic and environmental factors influencing health in these understudied settings.

Here we describe the overall structure of the CCPM Biobank and present initial findings supporting its simultaneous utility for research and clinical care transformation with over 200,000 enrolled participants and 33,674 genotyped participants as of March 2022. We characterize the current base of consented individuals and our first cohort of genotyped participants. We demonstrate the utility for research via association studies for ten common cardiovascular, immune, metabolic, and anthropometric polygenic traits. Finally, we describe the opportunities to translate research insights directly into clinical applications, including return of actionable clinical genetic test results as we realize the Anschutz Medical Campus and UCHealth vision for personalized medicine for all.

## METHODS

### CCPM Planning and Development

In 2014 the major partners at the Anschutz Medical Campus—University of Colorado Anschutz Medical Campus, UCHealth, Children’s Hospital Colorado and CU Medicine—came together to create the CCPM. At its inception, CCPM launched a campus-wide survey of needs and interests in personalized medicine present in both the university and health system for both clinical implementation and research. CCPM members, in discussions with the Colorado Multiple Institutional Review Board (COMIRB) with input from UCHealth Patient and Family Advisory Councils, began developing a novel consent model that supported the dual goals of the CCPM Biobank and held focus groups to develop consenting materials that balanced scientific accuracy and approachability.^5^

### Participant Recruitment and Engagement

The CCPM Biobank began enrollment in September 2015 with a self-consent, paper-based, process for adult participants (age ≧18 years, who could consent for themselves in English) receiving medical care at UCHealth University of Colorado Hospital. Children were not enrolled. The two page consent form authorized collection of blood samples leftover after clinical testing at UCHealth and provided broad consent for research and participant recontact, leaving open the opportunities for return of clinical genetic test results. Supporting information for consent included clinician education, patient information brochures, a frequently asked questions (FAQ) sheet, and a telephone helpline. This initial enrollment process was piloted within the Cardiac and Vascular Center at the UCHealth University of Colorado Hospital. In February 2016, the consent form was updated to allow for an additional tube of blood to be collected, dedicated to the Biobank. In July 2016, the consent process was expanded to include three additional clinics at the UCHealth University of Colorado Hospital, and to allow for consent via the UCHealth My Health Connection patient portal.

In October 2016, enrollment expanded again to additional UCHealth clinics in the Denver metropolitan area and we added an informational video to augment the online consent process.^6^ In August 2018, we transitioned entirely to an electronic self-consent model using My Health Connection, UCHealth’s online patient portal, and enrollment was opened to patients across the entire UCHealth system. To enable the return of clinical genetic test results to participants and their healthcare providers, in the Spring of 2018, we launched a pilot of return of results by recontacting CCPM Biobank participants with high-impact pathogenic variant findings. The consent form was also revised at this time to include additional information on clinical and research aims, sharing of data, Colorado legal statutes, and the Genetic Information Non-discrimination Act (GINA). In October 2018, we began a large secondary consent initiative to authorize actual return of clinical pharmacogenetic test results. In November 2019, we launched a revised consent form to include language allowing the CCPM Biobank to simultaneously return clinical genetic test results to participants and their healthcare providers in addition to performing research (referred to as the ‘unified’ consent).

To keep participants informed and involved in CCPM activities, we have implemented several engagement efforts including sending monthly on-boarding messages to welcome newly-enrolled CCPM Biobank participants and quarterly newsletters to all participants. Newsletters update participants on new partnerships, recent research activities within CCPM, and opportunities to become involved in CCPM Biobank activities, such as focus groups and new research studies performed in CCPM. Our website (www.cobiobank.org) is used to maintain participant engagement and to support educational efforts for both patients and providers around return of clinical genetic test results such as those related to pharmacogenetics, disease risk, and carrier status. Participant engagement approaches are in continual development due to the dynamic nature of the scope of the Biobank for research discovery and clinical care, and our continuously evolving capability with respect to return of clinical genetic test results.

### Sample Acquisition and Processing

Samples from early participants consisted of blood samples leftover after clinical testing at UCHealth clinical laboratories. The process of identifying consented participants and acquiring leftover specimens were manual processes and therefore resource- and time-intensive. To improve sample acquisition workflows, in conjunction with the February 2016 consent update we launched an automated sample collection process whereby participant consent recorded in the EHR triggers a blood collection order to collect a dedicated CCPM Biobank sample at the participant’s next clinical blood draw. This improved our collection of biospecimens, but there remained a significant gap between participants enrolled and samples collected, in part because not all active UCHealth patients would ever receive an order for a blood draw (e.g., ophthalmology). This was exacerbated by the onset of the COVID-19 pandemic. In March 2021 we piloted the collection of saliva samples at limited UCHealth locations during COVID-19 vaccination clinics for patients who consented to participate in the Biobank.

The CCPM Biobank Laboratory is College of American Pathologists (CAP)-accredited and Clinical Laboratory Improvement Amendments (CLIA)-certified for high-complexity testing. Samples are accessioned into a clinically-compliant Laboratory Information System (LIS) (SLIMS, Agilent, Inc.), which links patient demographics to samples and their derivatives. Samples collected in clinical labs, including blood (4mL, EDTA) and saliva (2mL, Genefi, IsoHelix), are transported to the CCPM Biobank Laboratory. Genomic DNA is extracted into 2D barcoded tubes in a 96-well plate format on a FlexSTAR+ instrument (Autogen Inc.) utilizing Flexigene chemistry (Qiagen). Nucleic acid yield is determined by fluorescence (QuantiFluor ds DNA system, Promega) and UV absorbance, and these values, along with characteristics such as volume and storage location, are linked to each sample in the LIS. DNA samples are stored at 4°C prior to genotyping and at −20°C for long-term storage.

Genotyping was performed on one of two different customized versions of Illumina’s Infinium Expanded Multi-Ethnic Genotyping Array (MEGA-EX) according to manufacturer’s instructions. We chose the this platform as the primary genotyping array given our extensive experience in development of genotyping platforms in The Consortium on Asthma among African-ancestry Populations in the Americas (CAAPA)^7,8^ and The Population Architecture using Genetics and Epidemiology (PAGE)^9–11^ studies and because it was designed with marker representation from numerous global populations to cover both clinical and scientific interests.^12^ Customized content on the MEGA-EX was composed of probes for >40,000 SNPs requested by University of Colorado Anschutz Medical Campus research community, with prior extensive QC on call rates and to ensure sample consistency (see Supplemental Methods). Among the ∼2.1 million genetic variants on MEGA-EX are several thousand with potential clinical relevance, a subset of which have either been analytically and clinically validated, or had a confirmatory assay developed, for later return to participants and their providers.

### Research Applications

In addition to genomic data, the CCPM Biobank contains rich phenotypic information through linkage to EHR. UCHealth has maintained a single Epic-based EHR system across all of its sites since 2011 and currently has records for over 8 million unique patients. Health Data Compass (HDC) (healthdatacompass.org), our institutional research data warehouse, extracts patient data from Epic including demographics, diagnosis codes, notes, flow sheets, lab values, admissions/discharges, and medications. Data are harmonized to the Observational Medical Outcomes Partnership (OMOP) common data model^13^ and linked to external data sources such as immunization records and vital statistics including death records from the Colorado Department of Health and Environment as well as Colorado All Payers Claims Database (APCD).^14^ These integrated data are stored in a HIPAA-compliant environment in Google Cloud. Approved researchers can securely access approved analytic data sets via Google BigQuery and secure cloud-based workspaces, allowing for easy access using Structured Query Language (SQL) or with Application Programing Interfaces (APIs) in R and Python. Data can be made available to researchers in identified or de-identified form and can be refreshed at regular intervals to support longitudinal research.

Oversight of proposed research using the CCPM Biobank resources is governed by the Access to Biobank Committee (ABC), composed of regulatory specialists, bioethicists, epidemiologists, informaticians, and statistical geneticists. In addition to regulatory approvals required by Health Data Compass (i.e., COMIRB approval for identified protocols, approval by UCHealth Privacy and Security Officers, etc.), the CCPM Biobank requires additional approval to ensure research requests respect availability of resources and are consistent with both the letter and spirit of the CCPM Biobank consent. The application process includes a description of the proposed research project, the analytical team, workflow, resource storage and security, participant inclusion and exclusion criteria, as well as a description of the project in lay language to allow participants to learn about active research projects through the CCPM Biobank newsletter. As of February 2022, ABC has delivered information on 38 *in silico* cohorts and approved 23 requests for data from CCPM researchers and external scientists.

### Clinical Applications

At present, two types of clinical genetic test results are being returned to CCPM Biobank participants and their providers when the participant has signed an appropriate consent: (1) high-impact pathogenic variants, and (2) pharmacogenetic results. We have implemented separate processes for returning these two types of results. Our decision to return high-impact pathogenic variants was taken with consideration of the policy from the American College of Medical Genetics and Genomics (ACMG)^15^ for reporting of secondary findings in clinical exome and genome sequencing, with a concentration on those genes for which actionable interventions exist to alter risk and/or disease outcome. Variants from MEGA genotyping that are interpreted to be pathogenic or likely pathogenic in high-impact genes of interest are confirmed in the CCPM Biobank Laboratory with a validated Sanger sequencing method. Due to the potential impact on participants and their families, our current return of results model for high-impact pathogenic variants includes an initial step of recontacting participants to confirm their consent and desire to receive such results. Once consent is confirmed, results are returned via telephone by a certified genetic counselor and placed in the EHR. Written information about their results is also made available to the participant, and participants are referred for clinical follow-up with specialists, as appropriate.

A summary of the CCPM Biobank clinical pharmacogenomics implementation process has been previously published.^16^ Briefly, the MEGA-EX array contains a number of known pathogenic and functional variants selected from ClinVar (www.ncbi.nlm.nih.gov/clinvar/),^17^ Online Mendelian Inheritance of Man (OMIM, omim.org),^18^ PharmGKB (pharmgkb.org),^19,20^ Clinical Pharmacogenetics Implementation Consortium (CPIC) (cpicpgx.org), and other databases of known variants that could be relevant to patient care. To streamline the pharmacogenomic implementation process, variants with clinical potential are validated on the array using complementary data generation approaches to ensure high fidelity and technical validity of results generated using the MEGA-EX platform directly rather than relying on confirmatory testing as in the case of high impact pathogenic variants. A custom, automated pipeline using BC Platforms (https://www.bcplatforms.com/) software provides the translation layer from a standard text format into a version that can be ingested by the UCHealth EHR system and filed as structured data elements. Structured results are then used to trigger drug-gene specific clinical decision support (CDS) tools (e.g., interruptive alerts, passive warnings), which provide clinicians with pertinent pharmacogenetic results and clinical recommendations at the point-of-prescribing.

### Provider Education

Provider education is an important part of our mission to facilitate personalized care. Due to well known limits in genomics knowledge among clinicians,^21–23^ local health professions’ groups and representative stakeholders were engaged throughout the development of our return of results process. Educational materials were created with involvement of relevant practitioner groups, including physicians, advanced practice providers, and pharmacists. These materials undergo at least annual assessment and revision to promote the use of genetic test results generated by the CCPM Biobank in clinical care. Evaluation is ongoing to ensure that these results are used effectively to positively impact patient diagnosis, screening, and treatment.

### CCPM Biobank Characterization and Genomic Replication

We identified all participants enrolled in the CCPM Biobank as of February 24, 2022 inclusive of those genotyped to calculate descriptive statistics on population demographics (gender, race/ethnicity, and age). Racial and gender identities with representation of less than 1% in the cohort were removed for privacy reasons. Median observation period was calculated using the entry dates of the first and last diagnosis codes in each patient’s record, excluding medical history diagnoses which are associated with prior dates of presentation, not the date entered in the record. The frequency and coverage (i.e., the percent of patients with one or more entries) of different data types (encounters, diagnoses, procedures, flowsheets, medications, laboratory tests) were calculated for the entire CCPM Biobank population from January 1, 2011 (the time of EHR implementation) to February 24, 2022. We also analyzed the prevalence of medical conditions in the CCPM Biobank using over 1200 phecode-based phenotypes^24^ derived from ICD9-CM and ICD10-CM codes using the PheWAS R package.^25,26^ Finally, maps of participant location were created using 3-digit zip code (as required by HIPAA de-identification regulations) and zip code tabulation area (ZCTA) from the United States Census Bureau.^27^ CCPM Biobank participant data were aggregated over the resulting regions and censored to remove 17 ZCTAs with a population of 20,000 or fewer persons based on census data from the most recent 3-digit count in 2000.^28^ All analyses were conducted using R version 3.6.0^29^ and a variety of packages for data processing, graphics, and reporting.^30–44^

We performed association tests at ten well-established loci in the GWAS catalog using imputed genotypes in the first set of 33,674 patients with MEGA genotype data (“Research Pilot”). We conducted extensive quality control on genotype batch, inter-plate concordance, and SNP and sample filtering on sites directly genotyped on MEGA (see Supplementary Methods), then imputed additional genotypes using the TOPMed Imputation Server^45,46^ in two batches, keeping the intersection of SNPs in both batches with r^2^ > 0.7 (see Supplemental Methods). Each association test used genotype, sex, age, the first five kinship-adjusted principal components by GENESIS^47–50^ as fixed effects, along with the genomic background correction (for highly polygenic traits and relatedness) present in REGENIE.^51^ We assessed proper calibration of association tests via the genomic control inflation factor lambda^52^ and standard qq plots. We then compared the odds ratios of the significant associations found within the CCPM Research Pilot study to previously reported findings in the GWAS catalog^53^ (accession numbers available in Supplementary Material) for each phenotype. To ensure consistent stranding, we ignored variants with ambiguous or unreported effect alleles. Confidence intervals were reported using a normal approximation to the effect size estimate to calculate the standard error.

## RESULTS

### Enrollment, Collection, and Participant History

As of March 2022, 200,673 participants have enrolled in the CCPM Biobank and 103,600 have provided a blood (n=100,542) or saliva sample (n=3,058) (Figure 1). Enrollment increased significantly starting in August 2018 when we moved to an electronic consent process that became available to patients across the entire UCHealth system that represents approximately 2 million active patients. Since that time an average of over 3000 participants enroll in the CCPM Biobank every month and an average of over 1700 samples are collected. The ratio of consent to samples (∼2:1) remains fairly consistent over the past several years and reflects the delay in sample collection inherent in our process to collect an additional tube of blood at a future clinical blood collection. About 60% of all participants have signed a consent (secondary consent or unified consent) to opt in to receiving clinical genetic test results from the CCPM Biobank (see Participant Recruitment and Engagement in Methods). In terms of retention, the median time of enrollment is 33.5 months; 726 participants have withdrawn from the study, and 6,398 have died.

**Figure 1.**
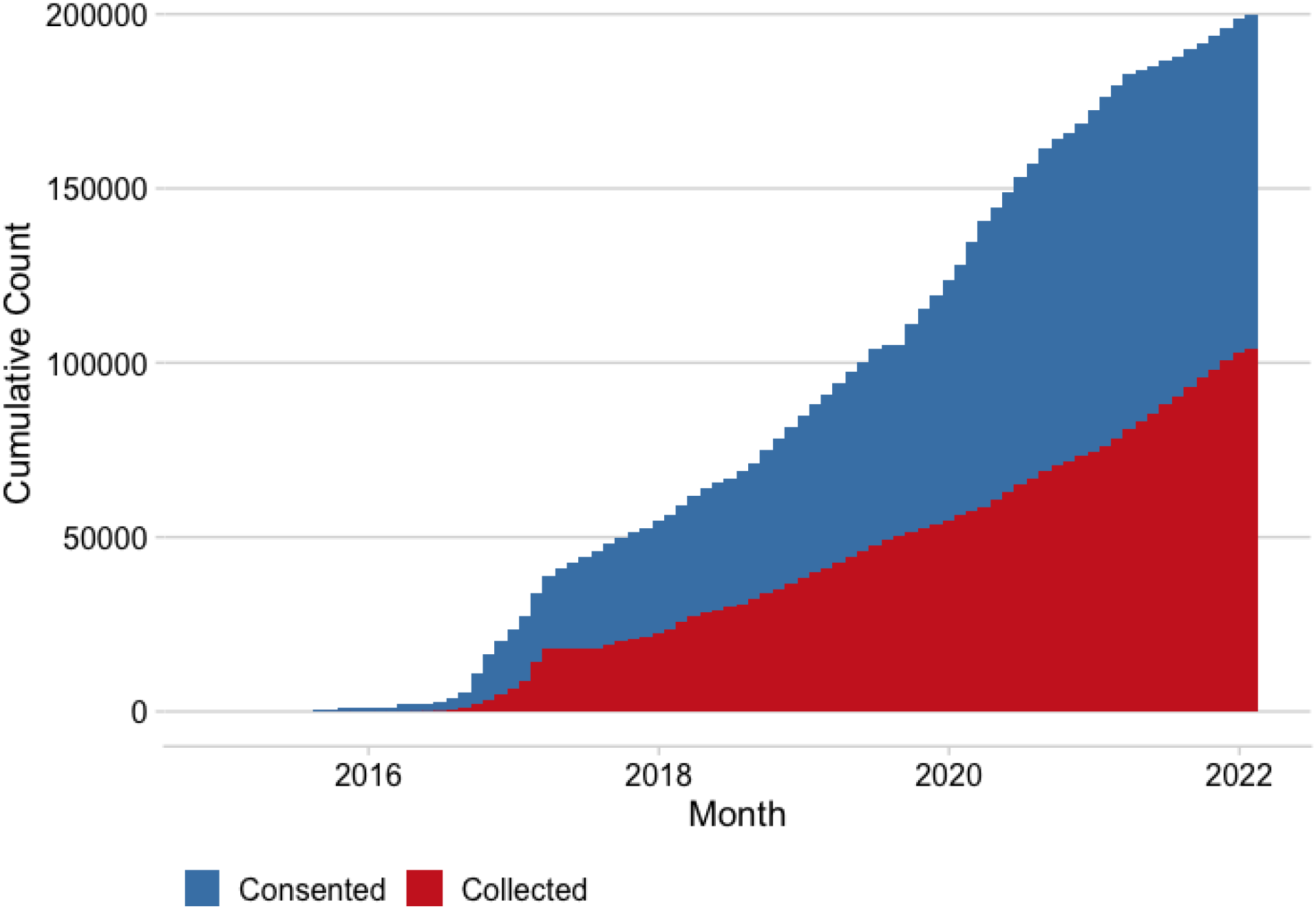
Cumulative enrollment (blue) and sample collection (red) of CCPM Biobank participants over time.

CCPM Biobank participants live in all 50 states as well as the District of Columbia (Figure 2A), although the majority (∼67%) reside along the Front Range region of Colorado, which includes the Denver Metropolitan Area, Fort Collins, and Colorado Springs (Figure 2B). Participants are predominantly female (59%), white (85%), and non-Hispanic (88%) reflecting the demographics of the underlying UCHealth patient population. The age of participants is mostly uniformly distributed for age groups from 30 to 69, and is somewhat lower for younger adults (18-29) and those aged 70 and older (Table 1).

**Table 1.**
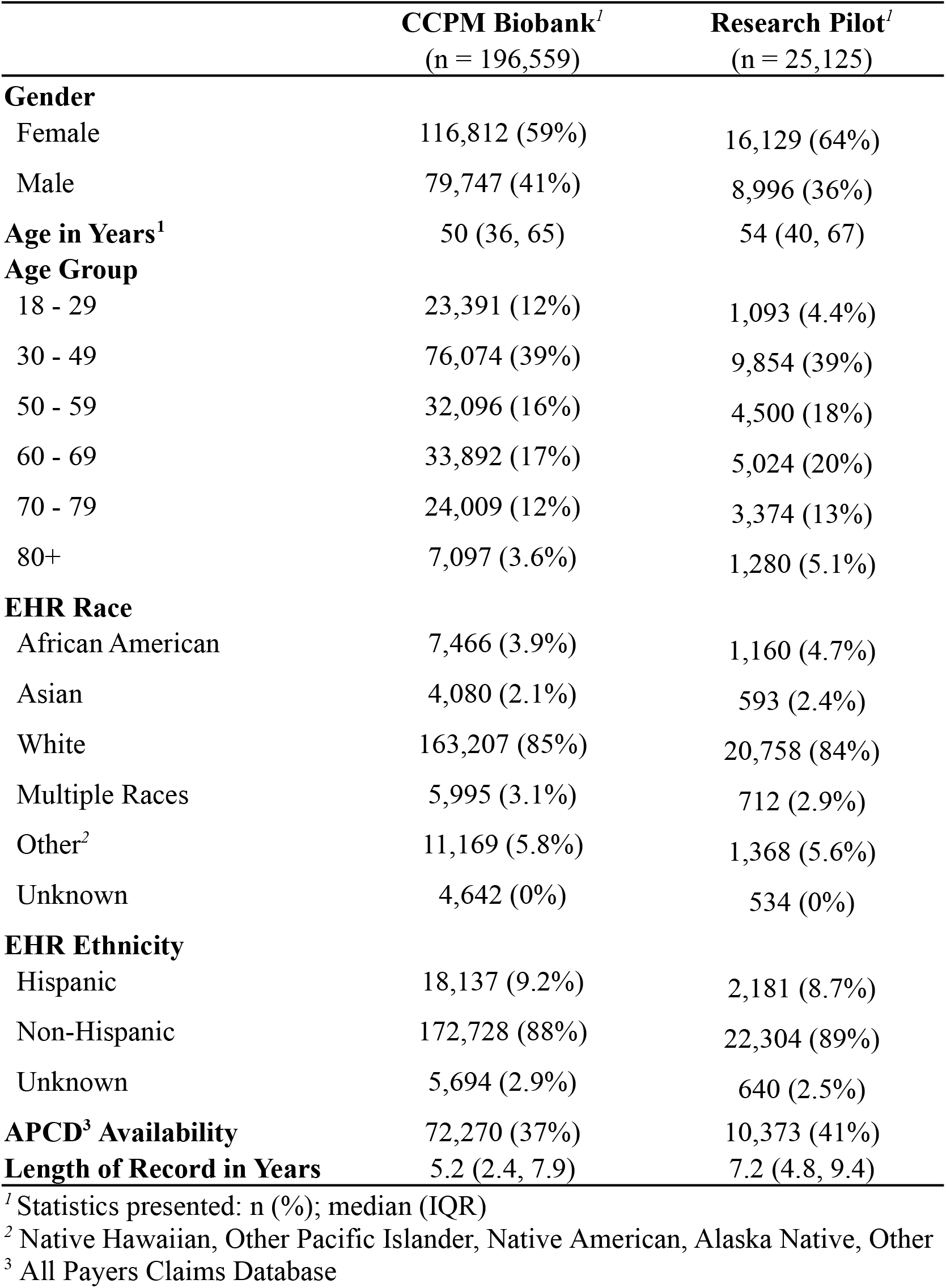
CCPM Biobank Participants and Research Pilot Demographics.

**Figure 2.**
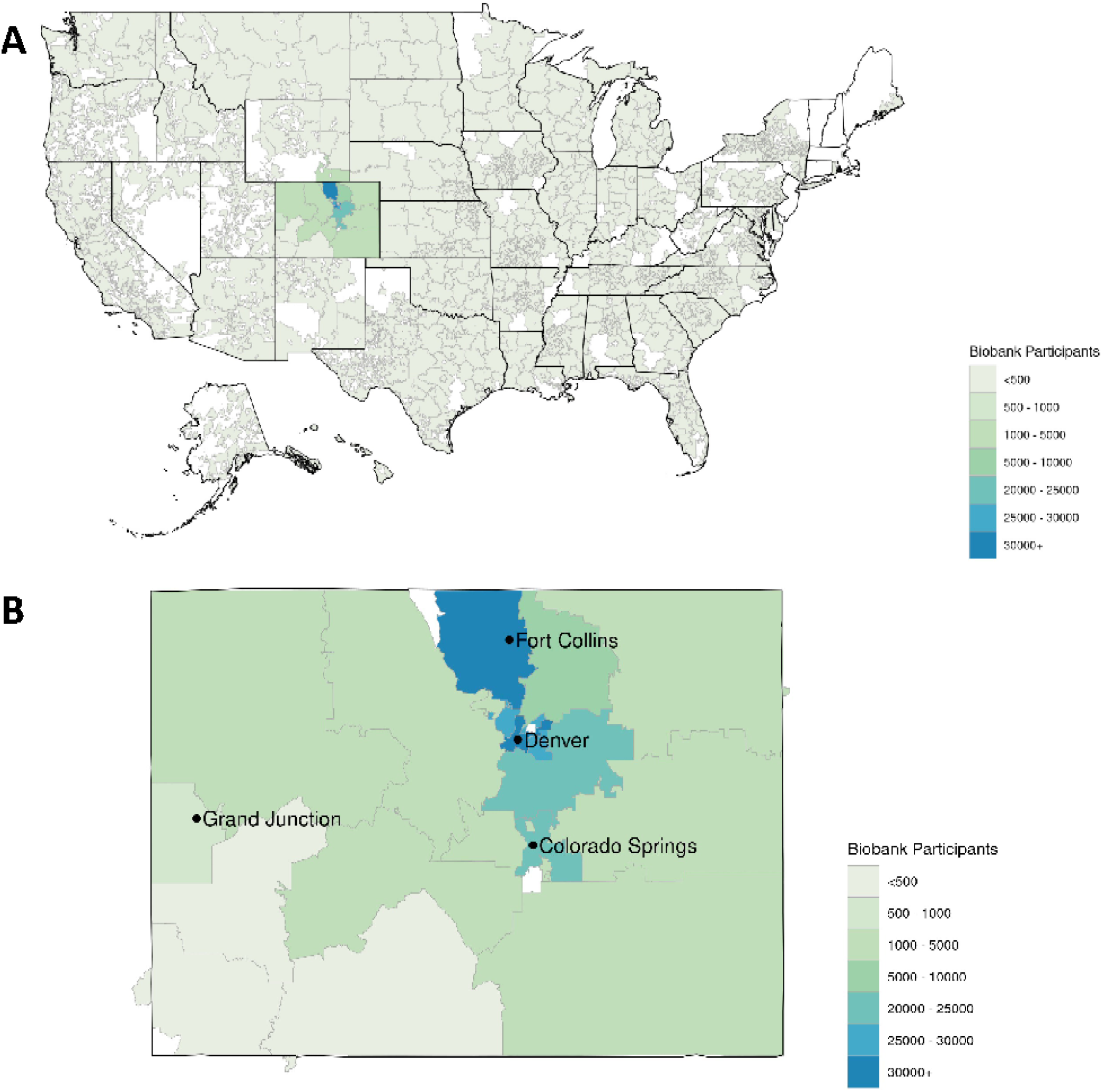
Density of recruitment of CCPM Biobank participants across A) the USA and B) Colorado, at the level of 3-digit zip codes.

Rich clinical history for each CCPM Biobank participant is available in the form of over 600 million data points in the EHR. Across a median observation period of 5.2 years, more than 99% of participants have information for at least one clinical encounter, diagnosis codes, and procedure codes (Table 2). We used ICD-9-CM and ICD-10-CM diagnosis codes in the EHR to derive more than 1200 phecodes covering a wide range of complex diseases for each participant. Nearly 50% of participants have a phecode for an endocrine/metabolic disorder, largely driven by disorders in lipid metabolism (Figure 3). Nearly 25% of participants have the phecode for hypertension, which is the single most frequent disorder noted in our population (Figure 3).

**Table 2.** Clinical Data Availability for CCPM Biobank Participants

| **Event Type** | **Total Event Count** | **Participant Count**<br>**(% cohort**<br>**represented)** |
| --- | --- | --- |
| Encounter Entry | 40,052,867 | 196,370 (99.9%) |
| Diagnosis Code | 40,576,473 | 194,554 (99.0%) |
| Procedure Code | 125,637,476 | 196,236 (99.8%) |
| Flowsheet Entry | 502,664,992 | 195,313 (99.4%) |
| Medication<br>Order | 18,583,715 | 181,989 (92.6%) |
| Laboratory Value | 85,632,533 | 183,352 (93.3%) |

**Figure 3.**
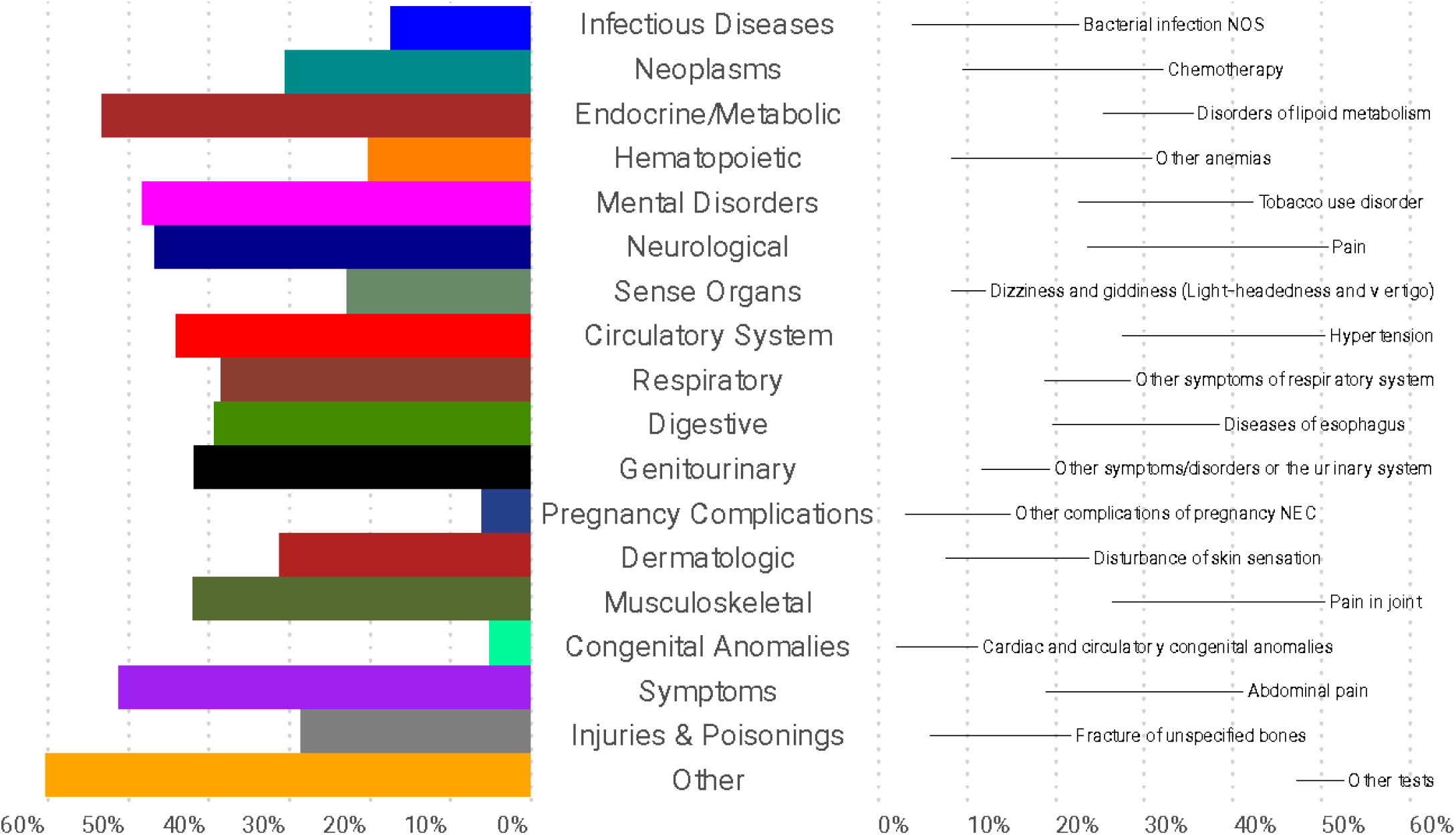
CCPM Biobank participant proportional phecode use by domain (left) and specific phecode (right) as derived from the EHR across participants in the entire CCPM Biobank

### Genotyping and Imputation

We chose the first 34,435 CCPM Biobank participants with verified consents to be genotyped on our custom MEGA^12^ platform, which contains ∼2 million markers selected for representation across diverse populations. Extensive quality control metrics were used to filter markers and samples (see Supplemental Methods). The genotyping success rate exceeded 98% of samples, and sex concordance between inferred genetic sex and sex or gender as reported in the EHR was >99.8%; of the 75 sex discrepancies, only 8 were unable to be resolved by EHR review for sex chromosome abnormality or transgender status. After QC was complete, 33,674 participants genotyped at 1,696,932 sites remained. We leveraged these sites to perform whole genome imputation using the TOPMed reference panel to yield over 98 million loci. After filtering to loci with confident imputation quality, ∼50M sites remained (see Supplemental Methods).

### Population Structure and Relatedness

Although family relationships often are not documented in EHR data, precise family structures can be inferred using dense genotype information. To infer familial relationships, we used our PONDEROSA algorithm,^54^ a method robust to arbitrary levels of endogamy, and KING-robust.^50^ We identified 761 first-degree relative pairs (comprising both parent-offspring and sibling pairs), 300 second-degree relative pairs, and 465 third-degree relative pairs (Figure 4A). From these relatives, we reconstructed numerous pedigrees, the largest of which is composed of 5 individuals and spans 3 generations.

**Figure 4.**
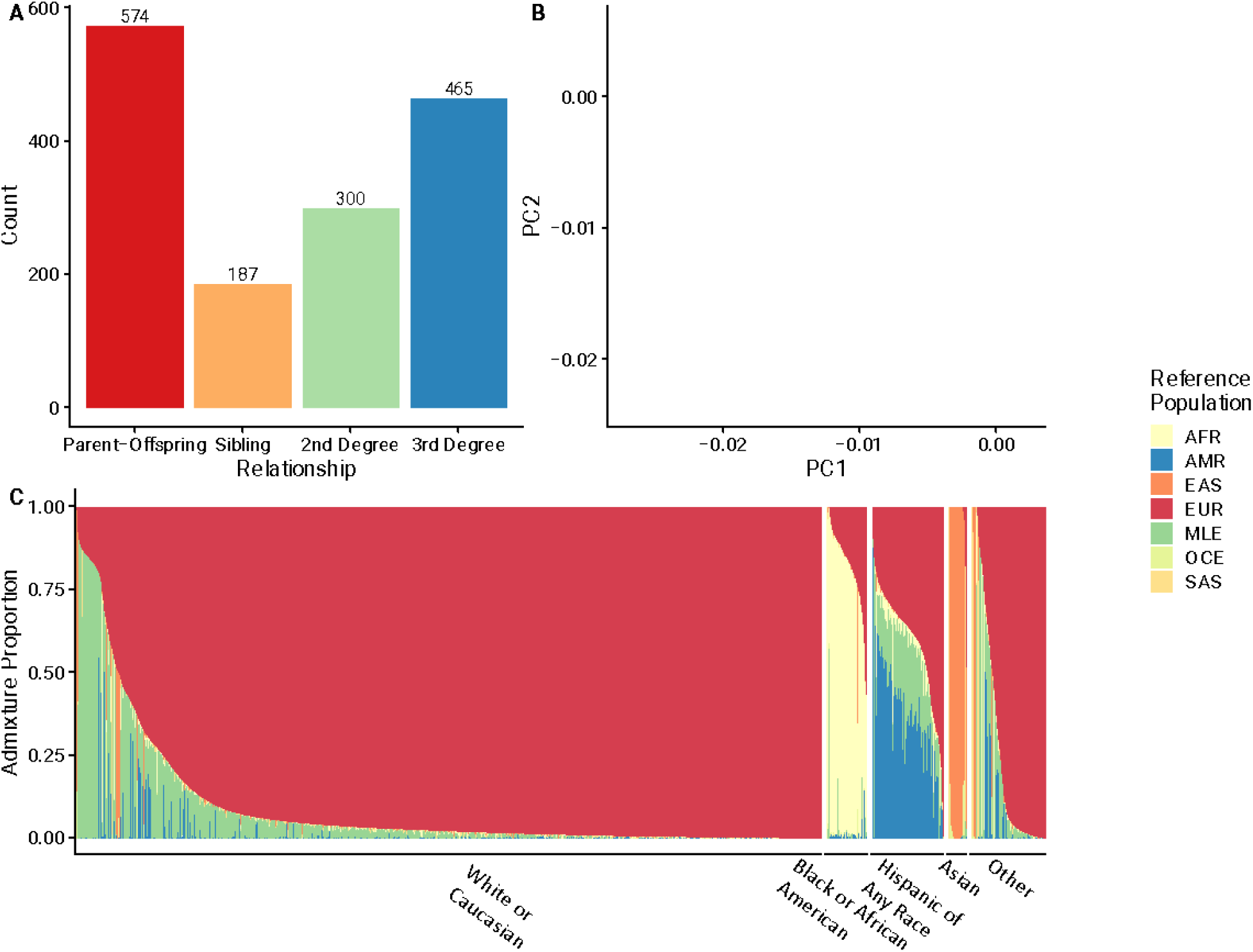
Ancestry and Population Structure estimates in the CCPM Biobank. A) Counts of closely-related pairs of individuals as determined by PONDEROSA. B) Principal Components Analysis of CCPM Biobank participants (gray points) overlaid with individuals from a global reference panel (colored points), and C) Admixture estimates for CCPM Biobank participants from the five largest EHR race/ethnicity categories. 1000 Genomes and Human Genome Diversity Project (HGDP) data are provided for reference.

Race and ethnicity information encoded in the EHR in the form of census categories is available for each Biobank participant, however ambiguities in racial and ethnic categories or mis-classification of race/ethnicity during clinical visits ma1y result in classifications that are inappropriate in genetic analyses (e.g., when controlling for population structure). We used genome-wide SNP data with different dimension reduction methods (see Supplemental Methods) to infer genetic ancestry cluster memberships for each CCPM Biobank participant. Although the majority (∼80%) of CCPM Biobank participants are of primarily European descent, participants represent a wide spectrum of diverse ancestries (Figures 4B-C), including substantial populations with genetic ancestry from Africa, South Asia, East Asia, and the Americas. The full spectrum of ancestry is further highlighted with global ancestry proportions (Figure 4C).

### GWAS Catalog Replication Findings

To evaluate the utility of the CCPM Biobank as a research tool, we conducted genetic association tests on 10 traits with previously well-established causal loci including Alzheimer’s disease, rheumatoid arthritis, psoriasis, multiple sclerosis, hypothyroidism, type 1 and type 2 diabetes, obesity, breast cancer, and asthma (see Pilot Analyses in Methods and Supplemental Methods), and compared the effect sizes of the most significant SNP for each trait with overlapping associations previously reported in the GWAS catalog.^53^ The effect sizes of overlapping SNPs are largely consistent with those previously reported (Figure 5), with the most significant SNPs having effect sizes in the same direction and general magnitude as reported. We note that although most of these associations would be significant after correction for multiple testing (as in a genomewide association study, for example), two SNPs associated with asthma and breast cancer would not. We recognize that our power to replicate previously-validated SNPs is limited in some instances by low case counts, especially when considering SNPs with modest effect sizes, as is consistent with other non-ascertained biobank cohorts.

**Figure 5.**
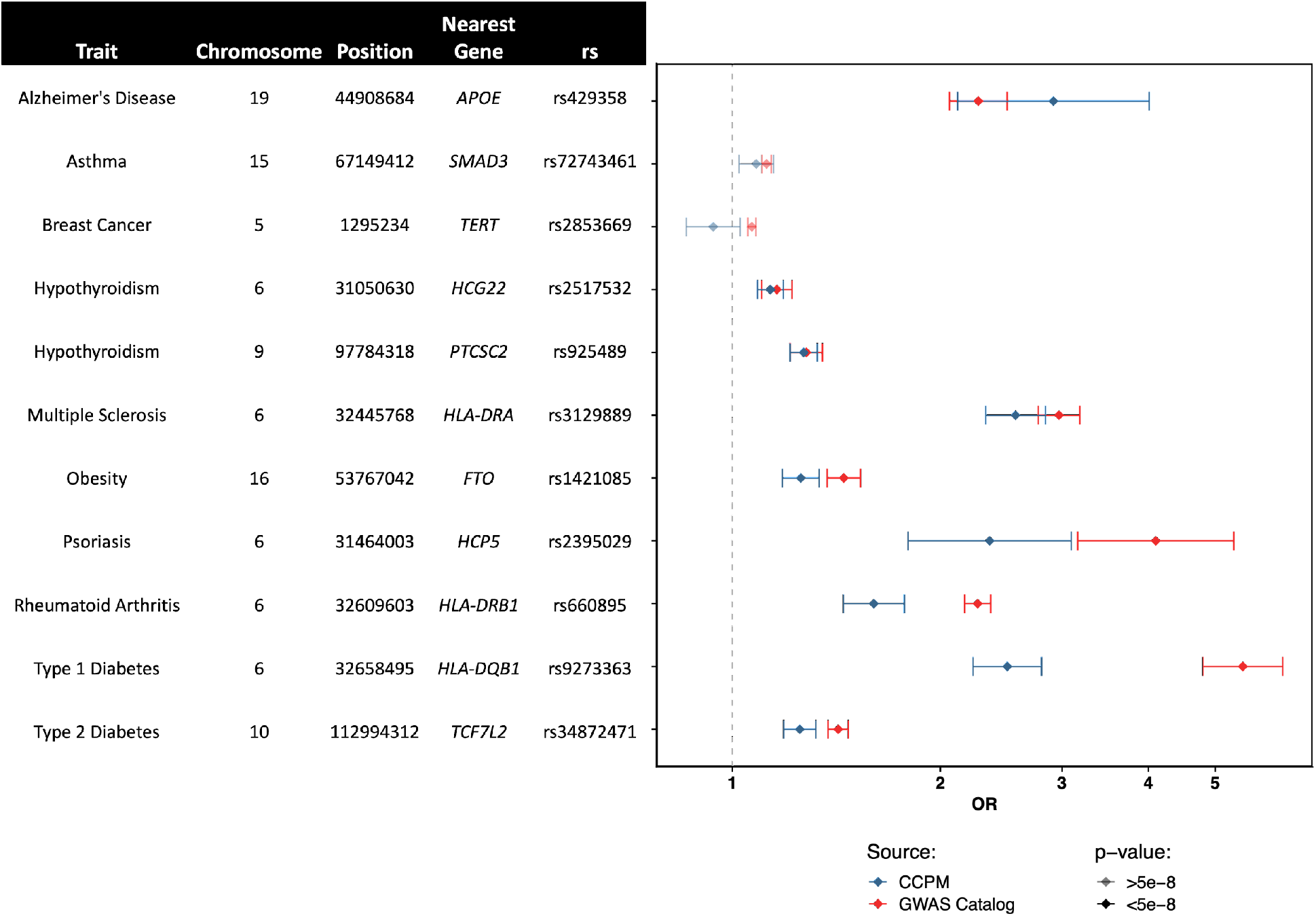
Replication of known hits in the CCPM Biobank across a range of traits, comparing CCPM Biobank findings with REGENIE to those found in the GWAS Catalog. For all, the risk-increasing allele is compared, and with multiple reporters, the largest dataset in the GWAS Catalog was used for reference.

### Clinical Applications

Nearly 10,000 of the 33,674 genotyped participants have signed a consent to receive clinical genetic test results. Currently, these results include high-impact pathogenic variants (HIPV) associated with increased risk for cancer, cardiac conditions and other rare genetic conditions (expected to grow to ∼1-2% of participants with the current HIPV list) and those that predict response to certain medications (expected to grow to nearly 100% of participants).

As of May 1, 2022, we have returned HIPV findings to 25 participants and their providers as part of a pilot to return these results to participants (manuscript in preparation). Six participants have variants in genes that confer increased cancer risk (i.e. hereditary breast and ovarian cancer and Lynch syndrome), 14 participants have mutations in genes that confer increased risk for cardiac-related conditions (e.g. cardiomyopathy, familial hypercholesterolemia), and one participant has a rare genetic condition. Feedback from our first 25 participants who have received these results has been very positive (manuscript in preparation). Most of the participants who were referred for clinical follow-up have either had or scheduled appointments with specialty providers and/or their primary care provider, and the majority have shared their results with family members. We continue to return these types of results as more participants opt to update their CCPM Biobank consent and become eligible to receive clinical genetic test results.

In 2019, we first preemptively returned *CYP2C19* pharmacogenetic results (i.e., *2, *3, *17) to the EHR via a custom, automated pipeline (see Methods) and we released the first CDS tool for clopidogrel. As of April 1, 2022, we have preemptively returned *CYP2C19* and/or *SLCO1B1* clinical pharmacogenetic results to 4,133 CCPM Biobank participants and their providers (see Methods). CDS tools are currently in operation for ten medications affected by *CYP2C19* variants [i.e., clopidogrel, voriconazole, citalopram, escitalopram, proton pump inhibitors (dexlansoprazole, lansoprazole, omeprazole, pantoprazole), brivaracetam, and clobazam] and seven statin medications affected by *SLCO1B1* variation (i.e., atorvastatin, fluvastatin, lovastatin, pitavastatin, pravastatin, rosuvastatin, simvastatin). To date, 420 drug-gene interaction alerts have been triggered for 378 CCPM Biobank participants. The most common drug-gene interaction alerts are for proton pump inhibitors (54.5%), followed by statins (22.6%), es/citalopram (21.9%), and clopidogrel (1%). An in-depth analysis of the relationship between drug-gene interaction alerts and subsequent clinical actions is underway. We expect to begin returning results for the next pharmacogene, *DPYD*, in Summer 2022.

### Research Applications and Participant Engagement

The ability to recontact participants is a defining feature of the CCPM Biobank and a necessary component to CCPM’s mission. Responses to health-related surveys and questionnaires from participants provide an invaluable resource to researchers who desire information about participants that may not be readily available in the EHR. To this end, the CCPM Biobank administered surveys in 2020-2021 to ∼180,000 CCPM Biobank participants to gain information about their experiences during the COVID-19 pandemic including testing, symptoms, healthcare utilization, and impact on health and well-being. Over 25,000 participants responded to the surveys (14% response). These data are linked with clinical data from the EHR and available for additional research.^55^

Maintaining participant engagement via on-going outreach is critical for establishing trust and transparency, e.g. informing participants of new research and industry partners with whom we share data, facilitating recontact for research opportunities and return of results, and optimizing retention. Regular outreach also allows us to provide education to our participants around genomics and personalized medicine, and to showcase studies for which they have contributed. Email open rates for our newsletter and on-boarding messages hover around 30% and 60%, respectively.

## DISCUSSION

The dual research and clinical mission of the CCPM Biobank provides unique opportunities for researchers, clinicians, and participants to be involved in and contribute to translational research that informs healthcare. The CCPM Biobank resource presently contains clinical data from the EHR for over 200,000 participants and dense array genotyping for nearly 34,000 participants, and this resource continues to grow steadily. The convenience of our self-consent model via the Epic patient portal (My Health Connection) allows us to reach patients across the entire UCHealth system and the ability to tailor educational messages in both English and Spanish. Participants provide broad consent to use collected samples and associated EHR data in wide-ranging health related research, including various-omics domains. They also consent to future contact for other studies, which serves as an invaluable resource for new study recruitment. Over time, the depth and breadth of research that can be supported by the CCPM Biobank will only expand with continued participant enrollment and engagement, including through participant surveys.

Nearly 60% of our participants have provided consent for return of clinical genetic test results and this percentage will increase over time as more participants sign the unified consent for both research and return of results. Return of clinical genetic test results is made possible because the Biobank Laboratory is both CLIA-certified and CAP-accredited allowing for direct return of clinically actionable results without the need for orthogonal confirmatory testing or to collect a new clinical sample. Our consent allows for multiple methods to screen for high-impact pathogenic variants which when returned to participants provides opportunity for clinical intervention, including earlier screening, diagnosis, and treatment. Return of these results for use in the clinical setting provides an important pathway to improve health and responsibly advance the frontiers of personalized medicine. Beyond reporting these high-impact pathogenic variants, efforts to return results for pharmacogenetic variants that alter drug metabolism and/or transport are already underway,^16^ and future advances in genetics will present new opportunities to directly impact patient health. As advancements are made in gene therapies and other genomic technologies, variants with limited to no clinical evidence today may be returned at a future time when clinical utility has been demonstrated.

Research efforts in CCPM are supported by rich clinical data from the EHR, data from outside sources such as the Colorado All Payers Claims Database (APCD)^14^ and Colorado Department of Public Health and Environment (CDPHE), and the ability to include geocoded environmental information which are all integrated into the robust cloud-based health data warehouse. These resources, in addition to the use of standard data formats, such as the OMOP common data model (https://www.ohdsi.org/data-standardization/the-common-data-model/)^13^ used by the All of Us Research Program^56^ and other large initiatives, enable the CCPM Biobank to participate in collaborative science and contribute to other large-scale research efforts, such as the recently-assembled Global Biobank Meta-Analysis^57^ (globalbiobankmeta.org) and COVID-19 Host Genetics^58^ Initiatives.

CCPM’s framework of systematic genotyping with clinical and translational potential, combined with consent allowing for external partnerships, has allowed us to develop relationships with partners both in academia and industry. Notably, outside of our academic collaborations, we have developed large-scale partnerships including as the first health data warehouse in the cloud, and our various partnerships for generating data and providing clinical and translational implementation to aid in return of results at scale.

The genetic diversity in large scale biobanks and the CCPM Biobank is of increasing importance to fully understand population health disparities and improve personalized medicine applications within the Rocky Mountain region and elsewhere. In the CCPM Biobank, as is common elsewhere (e.g., Belbin et al.^59^), EHR Race/Ethnicity imperfectly recapitulates genetic ancestry and may be sub-optimal in many genetic analyses (e.g., when controlling for population structure). However, genetic ancestry analyses reveal a diverse spectrum of ancestries present in the CCPM Biobank that reflect much of worldwide diversity. In comparison to other biobanks,^60–62^ the CCPM Biobank has a substantial proportion of non-European ancestry populations, including ∼10% Hispanic individuals, ∼5% African American individuals, and other historically underrepresented populations in genomics and in population-based biobanks.^59–62^ Future research will involve finer-scale characterization of the ancestry of participants, particularly focusing on the unique population demographics of the Rocky Mountain Region.

CCPM Biobank participants come from diverse environments. With the intersection of geocoded data, we plan to monitor the health of the rural participants in the CCPM Biobank, which make up a substantial proportion of the total catchment in UCHealth as well as our participants. Rural health remains highly underrepresented in biomedical research,^63^ particularly in institutional biobanks concentrated in major metropolitan areas. The UCHealth catchment also contains areas with moderate and high altitude (including participants living >10,000 feet above sea level), which provides a unique context that would be rare to observe in most health systems in the United States.

Like most biobanks, there are limitations to phenotypes derived solely from billing codes and other discrete EHR fields, so we note that any and all associations identified here would be refined with proper computational phenotyping efforts. By combining aspects of notes, unstructured data, and individualized lab values in our health warehouse and within the EHR, we have developed a system for chart review through the Translational Informatics Service, the operational unit within CCPM that connects health data warehouse data with omics data. This can help inform future efforts including disease endotyping and reducing misclassification. Further, with our access to ongoing surveys with our participants, we hope to be able to access relevant covariate information as well as phenotypic outcomes that may not be present, or in limited scope, in traditional EHRs (e.g., sleep disorders, intensity and duration of symptoms during illness). Further, we recognize our electronic consent model has limitations in outreach to patient populations who may be less likely to interact with UCHealth via the web portal and/or who require consent materials in other languages. To address inequities in enrollment, we have started to work with community-based research groups to develop tailored materials and enhance our reach in under-represented patient populations. Nonetheless we must acknowledge that since spring 2020 we have seen growth in adoption of the patient portal particularly with the increase in telemedicine.

Our goal over the next few years is to continue to increase the number of participants with both clinical and genomic data in the CCPM Biobank into the hundreds of thousands, and to optimize return of clinically actionable results to eligible participants. We also strive to expand the reach and utility of our data assets internally and externally by collaborating with colleagues involved in global efforts such as the GBMI (gbmi.org) as well as disease domain-specific efforts such as the PAGE Study (pagestudy.org). Through CCPM-initiated efforts and collaborations with national and international partners facilitated by CCPM resources, we can discover novel insights to inform targeted personalized strategies to improve health outcomes.

## Data Availability

All data produced in the present study are available upon reasonable request to the authors.

## ACKNOWLEDGEMENTS

We are deeply indebted to the participants who have made the CCPM Biobank a reality, and financial support from UCHealth, Children’s Hospital Colorado (CHCO), CU Medicine, the Department and School of Medicine, and the Skaggs School of Pharmacy and Pharmaceutical Sciences. We especially acknowledge Elizabeth Concordia, Jena Hausmann, Donald Elliman, and John Reilly, David A. Schwartz, David Ross, Dan Theodorescu, Ann Thor, and the CCPM Governance Committee for their vision in launching a cross-institutional enterprise focused on delivering personalized medicine at the point of clinical care. The authors credit oversight and guidance from the CCPM Governance Committee, Advisory Committee, the entire Health Data Compass team, and past and present members of the Pharmacogenomics Implementation Committee Colorado (PICColo).

