## Supplemental Methods for "Building a Vertically-Integrated Genomic Learning Health System: The Colorado Center for Personalized Medicine Biobank"

### Customization of MEGA

The custom content of MEGA1 consists of input from 32 investigators (comprising 11,997 entries and 5,372 unique regions) who responded to a Request for Information (RFI) submitted to the Anschutz Medical Campus research community to seek faculty input about which SNPs, genes, or chromosomal regions should be on the MEGA-EX array. For genes or chromosomal regions selected, linkage disequilibrium (LD) tagging was performed at r2=0.8 for SNPs with minor allele frequency (MAF)>0.05 in 1000 genomes (TGP) and Illumina design score > 0.4, prioritizing SNPs already on the MEGA-EX backbone and SNPs with known functional annotation (damaging polyphen2, HVAR, or SIFT score for missense variants; CADD score > 20; or GERP++ > 2). Chromosomal regions and SNPs requested by ≥ 4 investigators had multiple probes designed by Illumina. There were 45,393 SNPs submitted to Illumina, of which 41,674 were added to the MEGA-EX.

The custom content of MEGA2 was built on the MEGA-Global backbone and consists of 68,425 total variants which including the following: all custom variants from MEGA1; 2,532 variants for newborn screening; 5,625 clinically pathogenic SNPs; 4,700 SNPs from PharmGKB; 19,538 SNPs from multiple studies within the GWAS catalog; 1,490 SNPs from the GWAS catalog with clinical significance; 6,458 loss of function variants found in 5 or more individuals in gnomAD; and 1,862 variants to help call copy number variants (CNVs) in clinically relevant CNV-rich regions and enhance coverage of Y and MT chromosomes.

### EHR data used for research QC

Study data were collected and managed using REDCap electronic data capture tools ^1^ hosted at the Institute of Translational Health Sciences (ITHS). REDCap (Research Electronic Data Capture) is a secure, web-based application designed to support data capture for research studies, providing: 1) an intuitive interface for validated data entry; 2) audit trails for tracking data manipulation and export procedures; 3) automated export procedures for seamless data downloads to common statistical packages; and 4) procedures for importing data from external sources.

REDCap at ITHS is supported by the National Center For Advancing Translational Sciences of the National Institutes of Health under Award Number UL1 TR002319.

Gender, ethnicity, and withdrawal status as of 5/15/20 for all Biobank subjects were provided via REDCap. Only patients with valid entries in REDCap were included as part of the final research release. No withdrawn patients were included in the REDCap release, and thus, were excluded from all analyses. Date of allogeneic bone marrow transplant (BMT) status and date of Biobank sample collection was provided from Health Data Compass. Any patient with an allogeneic BMT prior to Biobank sample collection was included in QC, but excluded from the final research release.

### Genotype aggregation

Genotyping for the data freeze was performed in two separate batches, each with a different customized version of MEGA (MEGA1 and MEGA2). The first batch consisted of 289 96-well plates run on the MEGA-EX platform comprising 25,817 unique CCPM samples and the second batch consisted of 96 96-well plates run on MEGA-Global platform comprising 8618 unique CCPM samples. For both batches, each plate included 6 control samples consisting of 1000 Genomes Project ^2^ (1000G) samples or Genome in a Bottle ^3^ (GIAB) samples that were used for plate and sample QC. Including all control samples, as well as duplicate GTC files called from both Autocall and Beeline, 36,745 GTC files were converted to single-sample vcf files using a modified version of Illumina’s GTCtoVCF code (<https://github.com/Illumina/GTCtoVCF>) with preference toward the GTC file called from Beeline where duplicates existed. Each vcf file maintained log R ratio, B allele frequency, call rate, as well as 44 internal controls that were used in GenomeStudio for QC. The single-sample vcf files were then converted to a multi-sample vcf file and then divided by plate using bcftools ^4^.

### Sample Quality Control

Samples were initially filtered on a plate-by-plate basis by checking for sex discordance, excess heterozygosity, high variant missingness, genetic duplicates, and control sample concordance. A plate was considered to have failed genotyping if > 5% of samples failed any of these tests within the plate. No plates failed.

Sex discordance checks were performed comparing sex provided by REDCap and sex inferred from genotype using PLINK (v1.9) ^5,6^. A sample is considered to be sex discordant if the inbreeding estimate on the X chromosome exceeded 0.3 for females or was less than 0.7 for males. There were 45 samples that were missing sex in REDCap, and an additional 154 samples that failed due to gender discrepancy.

We identified 274 samples that were missing > 5% of variant calls using PLINK (v1.9) ^5,6^ and were considered to have high missingness.

Tests for excess heterozygosity and genetic duplicates used a set of SNPs that were pruned for linkage disequilibrium using PLINK (v1.9) ^5,6^ to greedily prune variants in 50 SNP moving windows such that no pair of SNPs have an r^2^ > 0.5 (--indep-pairwise 50 5 0.5). Heterozygosity for each sample was calculated using PLINK (v1.9) ^5,6^, and samples with excess heterozygosity were those with inbreeding coefficients < -0.2. There were 43 samples with excess heterozygosity. We searched for duplicate samples using PLINK’s (v1.9) ^5,6^ ‘--genome’ command by looking for identity by descent pairs (IBD) with PI-HAT > 0.8. For these putative duplicate sample pairs, one sample from the pair was removed if the EHR confirmed that both had come from the same individual, and both samples of the pair were removed otherwise. We found 34 duplicate pairs; 26 had the same medical record number (MRN) (one sample sample was removed from each pair; 6 duplicate pairs were found between chips, and the MEGA2 result was chosen), 1 pair had different MRN’s but linked to the same individual (remove one sample), 4 had different MRN’s (removed both samples), and 3 pairs were suspected to be twins.

To check control sample concordance, we checked for allelic concordance of control samples on each plate using 308,981 SNPs that are present on MEGA-EX, 1000G ^2^, and GIAB ^3^, after removing strand-ambiguous SNPs, and ensuring that genotypes were all using the same strand. A control sample was considered discordant if > 0.1% of SNPs were discordant between the plate’s genotype calls and the available reference genotypes, and a plate was considered to have failed control sample concordance testing if > 1 control sample was discordant. No plates were removed due to failed control concordance testing.

### SNP Quality Control

SNP QC was performed separately by plate batch (MEGA1 or MEGA2). After removing SNPs due to array cluster failure, we removed SNPs with excess missingness, high differential in allele frequency between sexes, high differential in allele frequency by plate, high allelic discordance across 5 fixed control samples consistent across all plates, or not in Hardy-Weinberg equilibrium as described below. The total number of SNPs removed at each step is shown in Table 1.

**Table 1. Counts of SNPs removed at quality control steps for MEGA1 and MEGA2**

| **Quality control step** | **MEGA1** | **MEGA2** |
| --- | --- | --- |
| Cluster failure | 131070 | 12896 |
| Missigness | 3895 | 2201 |
| Differential allele frequency by sex | 85 | 3 |
| Differential allele frequency by plate | 285 | 0 |
| Allelic discordance | 2032 | 704 |
| Hardy-Weinberg | 12577 | 1550 |

Excess missingness was determined using PLINK (v1.9) ^5,6^. SNPs that were missing in more than 1% of samples were considered to have high missingness and were removed.

We checked for high differential in allele frequency between sexes by performing Fisher's exact test between males and females for each SNP using PLINK (v1.9) ^5,6^. SNPs with Fisher’s exact test p-value < 10^-12^ were removed.

We tested for high differential in allele frequency by plate by performing a Fisher’s exact test between the plate of interest and and all other plates, for all plates, for each SNP using PLINK (v1.9) ^5,6^. SNPs with Fisher’s exact test P-value <1e^-12^ on any plate were removed from the data.

Discordance across 5 consistent control samples across all plates was performed by calculating discordance (*d*) for each control at all markers according to $d = 1 - \frac{max(n_{AA}, n_{AB}, n_{BB})}{n_{AA} + n_{AB} + n_{BB}}$ where n_AA_, n_AB_, n_BB_ are the number of times the genotypes AA, AB, and BB are called for the individual at that marker as counted by PLINK (v1.9) ^5,6^. All SNPs with *d* > 0.05 were removed.

After removing any SNPs with excess missingness, high allele frequency differential by sex or plate, and high allelic discordance, we calcuated Hardy-Weinberg equilibrium for all SNPs in the three most frequent genetic ancestry groups in our dataset (Europeans, Latinos, and African Americans, see Inferring Genetic Ancestry below) using PLINK (v1.9) ^5,6^. SNPs with P < 10^-12^ were removed.

After combining MEGA1 and MEGA2 batches we performed QC with Fisher’s exact test using PLINK (v1.9) ^5,6^, and removed the 127 SNPs with P < 10^-12^, leaving 1,696,932 SNPs for analysis.

After removing a final 190 individuals due to withdrawing from CCPM, the final genotyped dataset includes 1,696,932 SNPS and 33,674 individuals.

### Inferring genetic ancestry

To infer genetic ancestry groups, we created a reference panel consisting of whole genome sequence data from unrelated individuals in the 1000G ^2^ and array data from unrelated individuals in the Human Genome Diversity Project (HGDP) ^7^. The reference panel consists of non-strand-ambiguous SNPs that are present in both 1000G and HGDP with MAF >= 0.05 and are in Hardy-Weinberg equilibrium.

We merged CCPM genotype data with the reference panel, ensured strand concordance, and filtered SNPs to keep those in Hardy-Weinberg equilibrium with MAF ≥ 0.05, and then pruned out SNPs that were not in linkage equilibrium using PLINK (version 1.9), ^5,6^ (--indep-pairwise 50 5 0.5).

We performed PCA on the merged dataset using PC-AiR in GENESIS ^8,9^, and reduced the first 5 PC’s to two dimensions using UMAP ^10^. We used the PCA-UMAP projection and k-nearest neighbors implemented in R package caret ^11^ with the number of neighbors set to 15 and the minimum distance set to 0.5. We used population labels in the reference panel (African, American, East Asian, European, Middle Eastern, Oceanian, South-Central Asian) to train the k-nearest neighbor classification.

### Imputation

SNPs were removed prior to imputation if they were strand-ambiguous (A<->T or C<->G) and the minor allele frequency compared to TOPMed reference was ambiguous (MAF for both Biobank and TOPMed > 0.3), and if they were monomorphic. Imputation was carried out on the TOPMed imputation server ^12,13^ with the array build of input on GRCh37/hg19, phased using Eagle v2.4 ^14^, then imputed with minimac4^15^ against the reference of TOPMed r2. A default allele frequency check against the TOPMed panel was included in the pipeline. Variants were imputed against the TOPMed Reference Panel ^12^ in separate batches (MEGA1 and MEGA2). Due to an upper limit of 25,000 sample size from the imputation server, we split MEGA1 individuals (N=25,362) to two equal sized inputs (N=12,681) for imputation and averaged the mean r^2^ values between the outputs per locus as the indication of imputation quality for MEGA1. We then removed variants with low imputation scores (r^2^ < 0.7) in either batch, and intersected the remaining variants between both batches to create the merged freeze. The final imputed data set consisted of 49,708,367 variants.

### Global Ancestry and Principal Components Analysis

Ancestry analyses were performed on 33,674 genotyped CCPM participants merged with genotype data from 1000G ^2^ and HGDP ^7^, and linkage pruned using plink’s (v1.9) ^5,6^ ‘--indep-pairwise’ command with parameters 10000 1000 0.1. Global ancestry proportions for each genotyped CCPM participant were determined with ADMIXTURE (v1.3) ^16^ using non-admixed continental-level populations (*k*=7, African, Indigenous American, East Asian, European, Middle Eastern, Oceanian, South-Central Asia) from 1000G and HGDP to supervise clusters. The final principal components analysis was performed with PC-AiR ^8,9^ using a kinship matrix derived from King’s (v2.2.7) ^9^ ‘--kinship’ to account for relatedness while calculating principal components.

### Inferring Relatives and Pedigrees

Counts for different degrees of relationships within CCPM participants were obtained from relationship inferences made by King’s (v2.2.7) ^9^ ‘--ibdseg’ command. The structure of pedigrees within CCPM were inferred using PONDEROSA ^17^.

### Trait Association Replication

We sought to replicate associations with ten traits at well-established loci found in the GWAS Catalog (v1.0.2, downloaded: 14 Oct 2021) ^18^. We used a custom R script to extract unique SNPS from the GWAS catalog for traits that map to well-defined phecodes and associations with p<10^-8^. We removed SNPs if a risk allele was not defined in the table,or if alleles at a position were strand-ambiguous (A/T or G/C), and selected the most significant association for duplicate SNPs within each trait. We then removed all SNPs except those that occur in well-established regions previously associated with each trait according to Table 2.

Phecodes for CCPM participants for the ten traits of interest (see Table 2) were inferred using the R package Phecode ^19,20^, which maps ICD-9 and ICD-10 codes from CCPM participants to phecodes. Association tests on imputed genotypes were performed using genome-wide regression analysis with REGENIE ^21^. A subset of 745,993 common genotyped variants (minor allele frequency >= 0.01, minor allele count >= 100) were used to build a whole genome model. The adjustment for unbalanced case/control ratio was made with approximate Firth regression. GWAS was run on up to 8 phenotypes simultaneously selected based on similar sample missingness. For each phenotype, inflation factor lambda was calculated, and q-q and Manhattan plots were generated.

To compare effect sizes between associations found within CCPM and the GWAS Catalog, we selected the most significant SNP from CCPM that overlapped a position with the filtered GWAS Catalog SNPs and had concordant alleles.

**Table 2. GWAS Catalog query information for association replication tests**

| Trait | GWAS Catalog Trait Query Term | Region | Chromosome | Query Start Position | Query Stop Position |
| --- | --- | --- | --- | --- | --- |
| Alzheimer’s Disease | “alzheimer's disease” | *APOE* | 19 | 44895796 | 44919393 |
| Asthma | “asthma” | *SMAD3* | 15 | 67063763 | 67195173 |
| Breast Cancer | “breast cancer” | 5p15 | 5 | 1 | 18400000 |
| Hypothyroidism | “hypothyroidism” | *HLA* | 6 | 28510120 | 33480577 |
|  |  | 9q22 | 9 | 87800000 | 99800000 |
|  |  | 22q12-q13 | 22 | 25500000 | 50818468 |
| Multiple Sclerosis | “multiple sclerosis” | *HLA* | 6 | 28510120 | 33480577 |
| Obesity | "overweight, obesity and other hyperalimentation", "obesity", "morbid obesity" | *FTO* | 16 | 53693963 | 54131941 |
| Psoriasis | “psoriasis” | *HLA* | 6 | 28510120 | 33480577 |
| Rheumatoid Arthritis | “rheumatoid arthritis” | *HLA* | 6 | 28510120 | 33480577 |
| Type I Diabetes | “type 1 diabetes” | *HLA* | 6 | 28510120 | 33480577 |
| Type II Diabetes | “type 2 diabetes” | *TCF7L2* | 10 | 112940247 | 113177678 |
